# Cross-reactive human antibody responses to H5N1 influenza virus neuraminidase are shaped by immune history

**DOI:** 10.1101/2025.09.02.25334929

**Authors:** Jordan T. Ort, Ashley Sobel Leonard, Shuk Hang Li, Reilly K. Atkinson, Lydia M. Mendoza, Marcos Costa Vieira, Sydney Gang, Sarah Cobey, Scott E. Hensley

**Affiliations:** Department of Microbiology, Perelman School of Medicine, University of Pennsylvania, Philadelphia, PA, USA; Division of Infectious Diseases, Children’s Hospital of Philadelphia, Philadelphia, PA, USA; Department of Ecology and Evolution, the University of Chicago, Chicago, IL, USA

## Abstract

H5N1 highly pathogenic avian influenza viruses have spread globally and pose a risk for a human pandemic. Prior studies suggest that early life exposures to group 1 influenza viruses (H1N1 and H2N2) prime antibodies that cross-react to the hemagglutinin of H5N1, which is also a group 1 virus. Less is known about how immune history affects antibody responses against the neuraminidase (NA) of H5N1 viruses. Here, we measured NA inhibition antibodies against multiple H5N1 viruses using sera from 155 individuals born between 1927 and 2016. We found that individuals primed in childhood with H1N1 viruses were more likely to possess higher levels of antibodies that cross-react with the NA of H5N1 viruses compared to individuals primed in childhood with H2N2 or H3N2 viruses. While young children rarely possessed cross-reactive NA antibodies, we found that childhood infections with contemporary H1N1, but not H3N2, viruses can elicit them. These data suggest that immune history greatly impacts the generation of cross-reactive NA antibodies that can inhibit H5N1 viruses.

## Main text

Highly pathogenic avian influenza viruses of the H5 2.3.4.4b clade continue to circulate widely in wild and domestic birds across the globe, with unprecedented transmission into a range of mammalian species since 2020^1–8^. In late 2023, B3.13 genotype H5N1 viruses spilled over into dairy cows in the United States, leading to widespread transmission within and between dairy farms^9–13^. In January 2025, the D1.1 genotype—a reassortant virus that acquired a divergent neuraminidase (NA) from endemic, low pathogenic avian influenza viruses in North America— also spilled over into dairy cattle in Nevada and Arizona^14,15^. As of June 2025, this outbreak has resulted in 41 infections in humans exposed to infected cattle^16^, highlighting the risk of H5 circulation within farmed animals. While circulating clade 2.3.4.4b viruses do not bind well to human-type receptors^17,18^ and there is no evidence for human-to-human transmission^2,19^, prolonged circulation in mammalian species may select for substitutions which confer enhanced binding, replication, and transmissibility in humans^20,21^. Pandemics can occur when influenza viruses—for which population-level immunity is low—acquire these abilities^22^.

Influenza virus exposures in childhood can form long-lived ‘immunological imprints’ that potentially provide protection against new pandemic strains encountered later in life^23,24^. Since 1918, there have been distinct periods of human circulation of different influenza virus subtypes; as a result, an individual’s birth year has a pronounced effect on the types of influenza viruses encountered early in life. H1N1 circulated widely from 1918–1957, reemerged in 1977, and then was displaced by a new, antigenically distinct H1N1 (sometimes referred to as H1N1(pdm09)) in 2009. H2N2 viruses circulated from 1957–1968, and H3N2 viruses emerged in 1968 and continue to circulate today.

It is well documented that H5N1 viruses typically cause more disease and mortality in younger individuals^25,26^, and this unusual age distribution of severe H5N1 infections is likely at least partially due to differences in immune history^27^. The hemagglutinin (HA) protein of influenza viruses can be separated into two general antigenic groups: group 1 (including H1, H2, and H5) and group 2 (including H3). We recently found that individuals ‘imprinted’ (i.e., initially infected) in childhood with H1N1 and H2N2 possess higher levels of antibodies that cross-react with the HA stalk domain of H5N1 viruses^28^. While these cross-reactive HA stalk antibodies likely contribute to protection against H5N1 viruses, immunity to NA may also be important in limiting disease severity^29,30^, particularly in the context of H5N1 viruses that share an NA subtype with human H1N1 viruses^31^. Previous studies have shown that some humans possess antibodies that cross-react to the NA of H5N1 viruses^32,33^; however, it is unclear if early-life infection history affects the likelihood an individual will possess these types of antibodies.

To assess pre-existing population-level N1 immunity, we first quantified NA inhibition (NAI) antibodies against H1N1(pdm09) virus (A/California/07/2009) in serum samples collected in 2017 from 155 individuals born between 1927 and 2016. Almost all adults had detectable NAI antibodies; however, titers were highest among the elderly and young adults, while those of middle-aged adults were generally lower (**Fig. 1a**). Many children had undetectable or low levels of NAI antibodies, suggesting limited prior exposure to H1N1 viruses at the time of sample collection. We next asked if the same samples contained antibodies capable of inhibiting the NAs of three H5N1 viruses: one historical virus from clade 1 (A/Vietnam/1203/2004) and two contemporary 2.3.4.4b viruses (A/Dairy Cow/Texas/24-008749-002-v/2024 [B3.13 genotype] and A/British Columbia/PHL-2032/2024 [D1.1 genotype]). We found that sera from most individuals possessed detectable cross-reactive NAI antibodies against H5N1 viruses, and that NAI antibody titers followed similar birth year trends as observed for H1N1(pdm09) (**Fig. 1b–d**). Although the magnitudes of H5N1 NAI titers were lower, we saw strong, positive correlations between these titers and those against H1N1(pdm09) (**Extended Data Fig. 1**; green), suggesting relatively high levels of antibody cross-reactivity across the N1 subtype.

**Figure 1.**
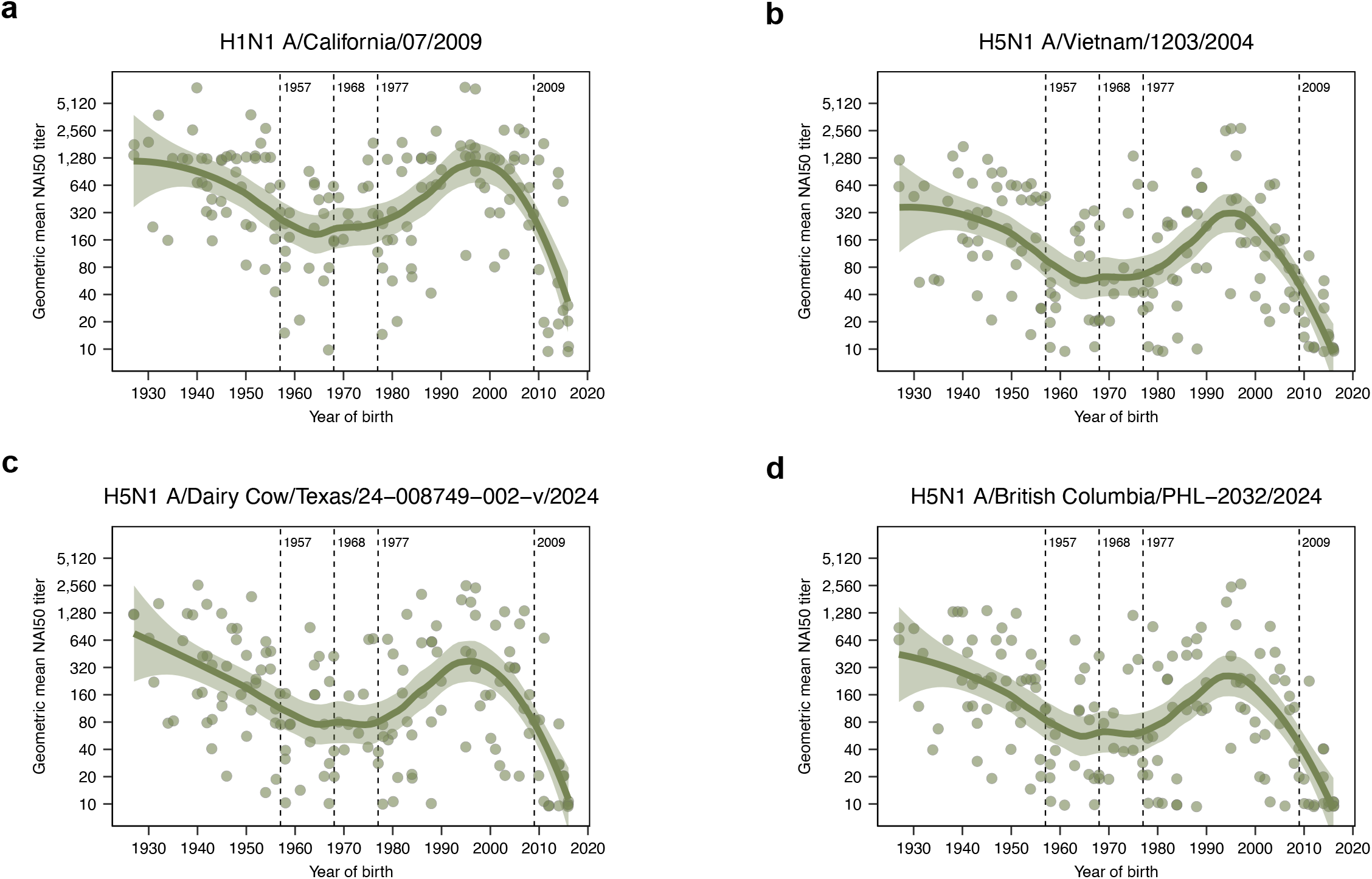
Imprinting with an N1 neuraminidase confers robust, cross-reactive N1 antibody responses. Serum samples, collected from healthy individuals at the Hospital of the University of Pennsylvania and the Children’s Hospital of Philadelphia (n = 155) in 2017, were assayed for NAI antibodies against H6Nx viruses expressing the NA of **(a)** H1N1 A/California/07/2009, **(b)** H5N1 A/Vietnam/1203/2005, **(c)** H5N1 A/Dairy Cow/Texas/24-008749-002-v/2024, and **(d)** H5N1 A/British Columbia/PHL-2032/2024. Vertical dashed lines represent the emergence of H2N2, H3N2, and H1N1, and H1N1(pdm09) in humans in 1957, 1968, 1977, and 2009, respectively. Each circle represents the geometric mean serum NAI50 titer for a single individual from two to three independent replicates, with locally estimated scatterplot smoothing curves (smoothing parameter = 0.5) and the associated 95% CIs.

Given that the highest N1 titers were seen in individuals born during periods of seasonal H1N1 circulation—from 1918 to 1957 and from 1977 to 2009—we asked if these results could be explained by the likelihood an individual was imprinted with an N1-containing virus. We found that N1 imprinting probabilities among adults were positively correlated with the NAI titers against each of the four tested N1 viruses, and that these had stronger Spearman correlations with titers than birth year (equivalent to age) or HA group 1 imprinting (**Extended Data Table 1**). We additionally computed bootstrap confidence intervals comparing the strength of correlations between NAI titers and each of these predictors. For each virus, we determined that N1 imprinting probabilities outperform both birth year and HA group 1 imprinting probabilities, while HA group 1 imprinting probabilities were not more strongly correlated with titers than birth year (**Extended Data Table 2**). As an alternative method, we also compared linear models that use birth year, N1 imprinting probability, or HA group 1 imprinting probability as a predictor for NAI titers. Against each virus, we found that models utilizing N1 imprinting probability outperformed the alternatives (ΔAkaike Information Criterion (AIC) values of 11.4–14.2 and 12.6–14.5 for HA group 1 imprinting probability and birth year, respectively; **Extended Data Table 3**). Altogether, these data support a role for immune imprinting in shaping cross-reactive N1 antibody levels in humans.

Previous work has shown that prior to H1N1(pdm09) emergence, only a subset of individuals possessed antibodies capable of inhibiting the NAs of a clade 2.3.4.4b H5N1 virus, with relatively low titers^32^. However, the oldest individuals in that study were born in 1950, providing limited information on the potential effects of pre-1957 H1N1 immune imprinting. We therefore assessed NAI antibody levels in adults using serum samples collected prior to 2009, with a focus on people born from 1918–1957. Using sera collected in 2005, we found high levels of pre-existing antibodies against the H1N1(pdm09) NA in older individuals, with decreasing titers as birth years approached 1957 (**Extended Data Fig. 2a**). Similarly, the oldest individuals possessed the highest levels of cross-reactive NAI antibodies against each of the tested H5N1 viruses (**Extended Data Fig 2b–d**). There were strong correlations between titers against H1N1(pdm09) and each of the H5N1 viruses (**Extended Data Fig. 1**; blue), suggesting that exposure to H1N1 viruses prior to 1957 elicited broad N1-directed antibodies. As we could not disentangle the contributions of age and of birth year in our previous analyses of samples collected in 2017, we reperformed the bootstrap analysis on a combined dataset using serum samples from both cohorts in this study (collected in 2005 and 2017), leveraging the 12-year gap in sample collection. Despite the relatively narrow year of birth range for the 2005 cohort— which may limit the ability to detect imprinting effects in this group—we found that N1 imprinting remained the most strongly correlated predictor of titers against each virus (**Extended Data Table 4**).

Finally, we completed a series of experiments to formally test the hypothesis that H1N1 infections induce antibodies that cross-react to the NAs of H5N1 viruses. We obtained serial sera samples from children infected with H1N1 during the 2023–24 and 2024–25 influenza seasons, with timepoints from both acute infection and convalescence. At acute timepoints, most children possessed low or undetectable levels of NAI antibodies against H1N1 viruses (A/California/07/2009 and A/Wisconsin/50/2022, a representative circulating strain) and B3.13 and D1.1 genotype H5N1 viruses (**Fig. 2a**). Following H1N1 infection, antibodies reactive to the N1s from H1N1 and H5N1 viruses increased significantly, whereas antibodies reactive to the N2 of an H3N2 virus (A/Thailand/8/2022) did not increase (**Fig. 2a**). We also measured antibodies in children infected with H3N2 viruses during the 2022–23 influenza season. We found that these children mounted NAI antibodies against the H3N2 virus but there was no significant rise in titer against any of the tested N1 viruses (**Fig. 2b**). Taken together, our data demonstrate that H1N1, but not H3N2, viruses elicit broadly reactive N1 antibodies, and that the low pre-existing titers seen in young children are likely due to a lack of or limited prior exposure to H1N1 viruses.

**Figure 2.**
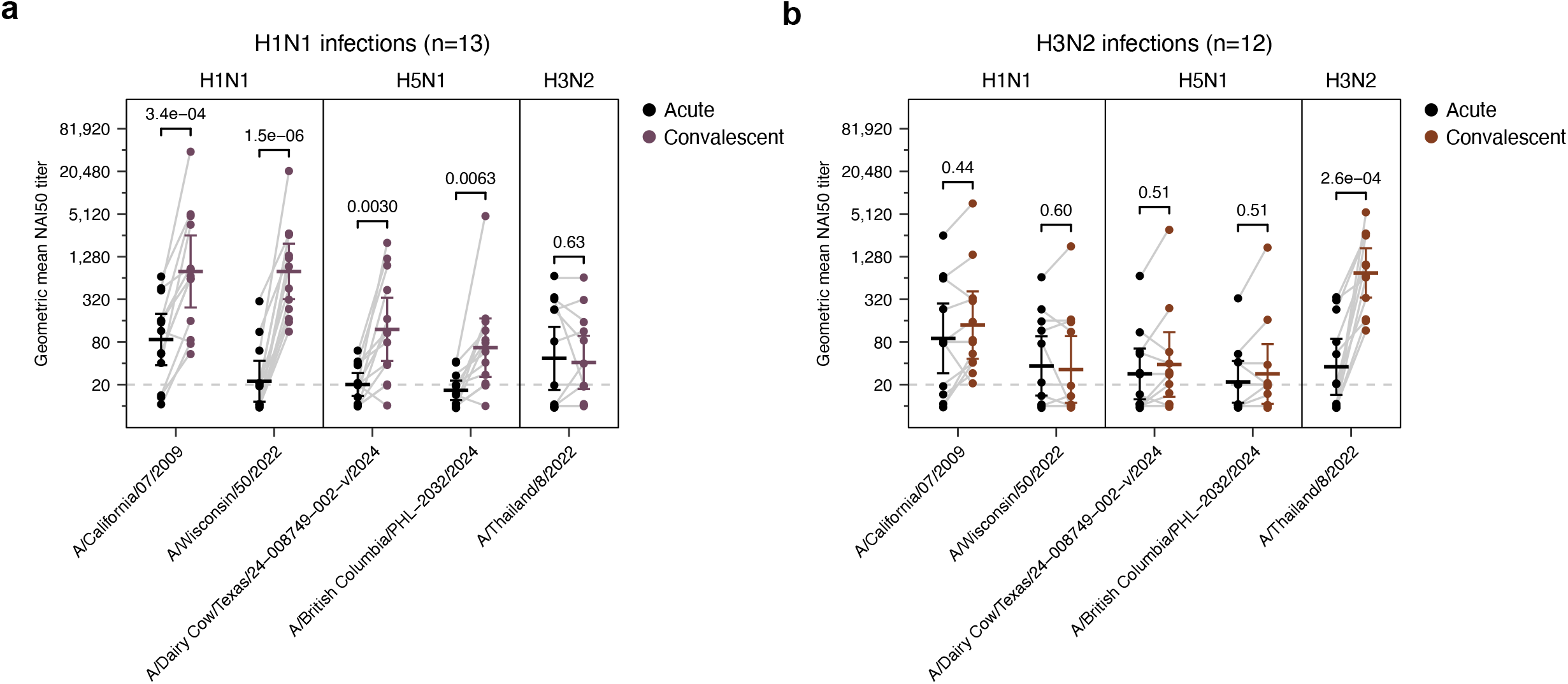
Contemporary H1N1, but not H3N2, infection elicits cross-reactive N1 antibodies in children. Serum samples were collected from children testing positive for H1N1 (n = 13) or H3N2 (n = 12) influenza A virus at the Children’s Hospital of Philadelphia during acute infection (within 7 days of symptom onset) and convalescence (more than 7 days since symptom onset). Sera from **(a)** H1N1-infected and **(b)** H3N2-infected children were assayed for NAI antibodies against H6Nx viruses expressing the NA of H1N1 A/California/07/2009, H1N1 A/Wisconsin/50/2022, H5N1 A/Dairy Cow/Texas/24-008749-002-v/2024, and H5N1 A/British Columbia/PHL-2032/2024. Each dot represents the geometric mean serum NAI50 titer for a single individual against a given strain. Acute titers are shown in black and maximum convalescent titers are shown in purple or red, with lines connecting. Geometric mean titers and the corresponding 95% CIs for each group are shown with error bars. Statistical differences between log_2_-transformed acute and convalescent titers were analyzed by two-sided paired t-tests, corrected for multiple comparisons with the Holm–Bonferroni method.

These studies indicate that immune imprinting affects the priming of NA antibody responses across the human population, which may impact differential H5N1 risks between age groups. Our data are consistent with recent work that suggest that in addition to HA group-level imprinting, homosubtypic NA imprinting can further modulate the risk of zoonotic influenza infection, including between H5N1 and H5N6 viruses which differ only by NA subtype^34^. This has far-reaching implications, as our studies predict that H5 viruses may cause differential levels of disease in the human population after reassortment with a novel NA.

While we found that initial exposure to an H1N1 virus confers high levels of cross-reactive N1 antibodies, and others have examined the role of NA imprinting in shaping age-specific seasonal influenza infection patterns^35,36^, further longitudinal studies are necessary to fully ascertain the importance and contribution of early-life infections. It is interesting to note that the peak in titers for people born during the 1990s cannot be explained by imprinting probabilities alone; higher titers in this birth cohort might be due to antigenic similarities between the NAs of 1990s seasonal H1N1 viruses and those used in our study, or potentially because many individuals in this cohort were primed with seasonal H1N1 in their childhood and then boosted with the antigenically distinct H1N1(pdm09) shortly thereafter. Testing if cross-reactive NA antibody responses could be elicited by immunization with an NA-based or NA-enriched vaccine may also prove useful, as conventional influenza vaccines do not have standardized NA contents^37^. It will also be important to utilize animal models to determine the protective capacity of cross-reactive NA antibodies. A recent study demonstrated that prior H1N1(pdm09) infections can protect ferrets from H5N1-induced mortality^38^; however, it remains unclear if this is mediated by HA or NA antibodies, T cells, or other means. Thus, passive transfer animal studies could clarify a potential protective role of cross-reactive N1 antibodies.

Although we identified similar patterns of cross-reactivity to each of the tested H5N1 NAs— including the reassorted NA of the D1.1 genotype—determining the epitopes targeted by these antibodies will be key to understanding their potential breadth. Given the diversity of N1 genes in wild birds^39^, and the propensity of H5 viruses to undergo reassortment^1,40^, it will be important to establish if certain N1 lineages could escape pre-existing immunity. A better understanding of how immune imprinting affects cross-reactive antibody responses within diverse human populations will be useful for risk assessment of new viral strains with pandemic potential.

## Data Availability

All data are shown in the main figures and the Extended Data. Source data are provided with this paper.

## Acknowledgements

We would like to thank the Penn Medicine BioBank and the Children’s Hospital of Philadelphia for providing sera samples from individuals of different birth years. We would also like to thank the NIAID and the VTEU clinical study teams from DMID 04-063 and DMID 04-076 for providing sera samples from clinical trials. This project was funded in part with federal funds from the National Institute of Allergy and Infectious Diseases, National Institutes of Health, Department of Health and Human Services, under contract no. 75N93021C00015 (S.E.H., S.C.) and grant number R01AI08686 (S.E.H.). S.E.H. holds an Investigators in the Pathogenesis of Infectious Disease Award from the Burroughs Wellcome Fund.

## Author Contributions Statement

J.T.O., L.M.M., A.S.L., and S.E.H. designed the experiments. J.T.O., S.H.L., R.K.A., and S.G. completed experiments and analyzed data. J.T.O. performed modeling studies, and M.C.V. and S.C. assisted with interpretation of these data. J.T.O. and S.E.H. wrote the manuscript and all authors contributed to editing the manuscript. S.E.H. supervised experiments and data analyses. S.E.H. obtained funding for the study.

## Competing Interests Statement

S.E.H. is a co-inventor on patents that describe the use of nucleoside-modified mRNA as a vaccine platform. S.E.H reports receiving consulting fees from Sanofi, Pfizer, Lumen, Novavax, and Merck. A.S.L. is now an employee of Sanofi but was affiliated with the Children’s Hospital of Pennsylvania when this work was completed. The authors declare no other competing interests.

## Notes

### Author Declarations

Experiments using human sera were conducted with the approval of the University of Pennsylvania Institutional Review Board and the Childrens Hospital of Philadelphia Institutional Review Board. Serological experiments were completed using de-identified samples. Informed consent was obtained for all individuals.

