## Extended Data for "Cross-reactive human antibody responses to H5N1 influenza virus neuraminidase are shaped by immune history"

**Extended Data Tables 1–4**
**Extended Data Figures 1–2**
**Online Methods**
**Extended Data References**

**Extended Data Table 1. Correlations between NAI titers against each virus and birth year, HA group 1 imprinting probabilities, and N1 imprinting probabilities.** We computed 95% confidence intervals using Fisher's transformation.

| NAI titers against | Correlate | Correlation Coefficient (95% CI) | *P* value |
|---|---|---|---|
| A/California/07/2009 | Birth year | 0.12 (-0.06,0.29) | 0.197 |
| | HA group 1 imprinting | 0.19 (0.01,0.36) | 0.035 |
| | N1 imprinting | 0.33 (0.16,0.48) | 2.81E-04 |
| A/Vietnam/1203/2004 | Birth year | 0.07 (-0.11,0.25) | 0.435 |
| | HA group 1 imprinting | 0.16 (-0.02,0.33) | 0.077 |
| | N1 imprinting | 0.35 (0.18,0.5) | 8.70E-05 |
| A/Dairy Cow/Texas/24-008749-002-v/2024 | Birth year | 0.1 (-0.08,0.27) | 0.296 |
| | HA group 1 imprinting | 0.19 (0.01,0.36) | 0.037 |
| | N1 imprinting | 0.32 (0.15,0.47) | 3.74E-04 |
| A/British Columbia/PHL-2032/2024 | Birth year | 0.12 (-0.06,0.29) | 0.189 |
| | HA group 1 imprinting | 0.22 (0.04,0.39) | 0.016 |
| | N1 imprinting | 0.35 (0.18,0.5) | 1.13E-04 |

**Extended Data Table 2. Bootstrap analysis comparing the strengths of the correlations between NAI titers against each virus and birth year, HA group 1 imprinting probabilities, and N1 imprinting probabilities.** Correlation coefficients are shown in absolute value. Boldface values indicate that predictor 1 had a stronger correlation with titers than predictor 2 did, based on the 95% bootstrap confidence interval.

| Virus | Predictor 1 | Predictor 2 | Predictor 1 titer correlation | Predictor 2 titer correlation | Observed difference | Bootstrap difference (95% CI) |
|---|---|---|---|---|---|---|
| A/California/07/2009 | N1 imprinting | Group 1 imprinting | 0.352 | 0.163 | 0.189 | **0.182 (0.058, 0.307)** |
| | N1 imprinting | Birth year | 0.352 | 0.072 | 0.280 | **0.249 (0.077, 0.385)** |
| | Group 1 imprinting | Birth year | 0.163 | 0.072 | 0.091 | 0.065 (-0.095, 0.172) |
| A/Vietnam/1203/2004 | N1 imprinting | Group 1 imprinting | 0.346 | 0.221 | 0.126 | **0.124 (0.004, 0.255)** |
| | N1 imprinting | Birth year | 0.346 | 0.121 | 0.225 | **0.212 (0.057, 0.364)** |
| | Group 1 imprinting | Birth year | 0.221 | 0.121 | 0.099 | 0.090 (-0.033, 0.203) |
| A/Dairy Cow/Texas/24-008749-002-v/2024 | N1 imprinting | Group 1 imprinting | 0.321 | 0.191 | 0.129 | **0.126 (0.007, 0.253)** |
| | N1 imprinting | Birth year | 0.321 | 0.097 | 0.224 | **0.206 (0.038, 0.361)** |
| | Group 1 imprinting | Birth year | 0.191 | 0.097 | 0.095 | 0.077 (-0.061, 0.189) |
| A/British Columbia/PHL-2032/2024 | N1 imprinting | Group 1 imprinting | 0.327 | 0.193 | 0.134 | **0.130 (0.003, 0.25)** |
| | N1 imprinting | Birth year | 0.327 | 0.119 | 0.208 | **0.197 (0.038, 0.344)** |
| | Group 1 imprinting | Birth year | 0.193 | 0.119 | 0.074 | 0.067 (-0.048, 0.173) |

**Extended Data Table 3. Comparison of linear models between NAI titers against each virus and birth year, HA group 1 imprinting probabilities, and N1 imprinting probabilities.** *k* indicates the number of parameters.

| Virus | Model | *k* | loglik | AIC | ∆AIC |
|---|---|---|---|---|---|
| A/California/07/2009 | N1 imprinting | 3 | -239.97 | 485.95 | 0 |
| | HA group 1 imprinting | 3 | -247.06 | 500.12 | 14.18 |
| | Birth year | 3 | -247.23 | 500.46 | 14.51 |
| A/Vietnam/1203/2004 | N1 imprinting | 3 | -248.3 | 502.61 | 0 |
| | HA group 1 imprinting | 3 | -254.63 | 515.27 | 12.66 |
| | Birth year | 3 | -255.46 | 516.91 | 14.3 |
| A/Dairy Cow/Texas/24-008749-002-v/2024 | N1 imprinting | 3 | -250.58 | 507.16 | 0 |
| | HA group 1 imprinting | 3 | -256.26 | 518.51 | 11.35 |
| | Birth year | 3 | -256.87 | 519.75 | 12.59 |
| A/British Columbia/PHL-2032/2024 | N1 imprinting | 3 | -250.38 | 506.76 | 0 |
| | HA group 1 imprinting | 3 | -256.36 | 518.72 | 11.95 |
| | Birth year | 3 | -256.85 | 519.7 | 12.94 |

**Extended Data Table 4. Bootstrap analysis comparing the strengths of the correlations between NAI titers against each virus and birth year, age, and N1 imprinting probabilities for combined data from 2005 and 2017 samples.** Correlation coefficients are shown in absolute value. Boldface values indicate that predictor 1 had a stronger correlation with titers than predictor 2 did, based on the 95% bootstrap confidence interval.

| Virus | Predictor 1 | Predictor 2 | Predictor 1 titer correlation | Predictor 2 titer correlation | Observed difference | Bootstrap difference (95% CI) |
|---|---|---|---|---|---|---|
| A/California/07/2009 | N1 imprinting | Age | 0.425 | 0.307 | 0.118 | **0.117 (0.027, 0.218)** |
| | N1 imprinting | Birth year | 0.425 | 0.288 | 0.137 | **0.135 (0.037, 0.235)** |
| | Age | Birth year | 0.307 | 0.288 | 0.019 | 0.019 (-0.017, 0.054) |
| A/Vietnam/1203/2004 | N1 imprinting | Age | 0.421 | 0.317 | 0.104 | **0.103 (0.002, 0.216)** |
| | N1 imprinting | Birth year | 0.421 | 0.320 | 0.101 | **0.102 (0.002, 0.214)** |
| | Age | Birth year | 0.317 | 0.320 | -0.003 | 0.004 (-0.037, 0.031) |
| A/Dairy Cow/Texas/24-008749-002-v/2024 | N1 imprinting | Age | 0.376 | 0.302 | 0.074 | 0.075 (-0.032, 0.186) |
| | N1 imprinting | Birth year | 0.376 | 0.250 | 0.126 | **0.129 (0.022, 0.236)** |
| | Age | Birth year | 0.302 | 0.250 | 0.052 | 0.052 (-0.02, 0.086) |
| A/British Columbia/PHL-2032/2024 | N1 imprinting | Age | 0.408 | 0.329 | 0.079 | 0.079 -0.024, 0.183) |
| | N1 imprinting | Birth year | 0.408 | 0.277 | 0.13 | **0.129 (0.025, 0.232)** |
| | Age | Birth year | 0.329 | 0.277 | 0.052 | **0.051 (0.019, 0.086)** |

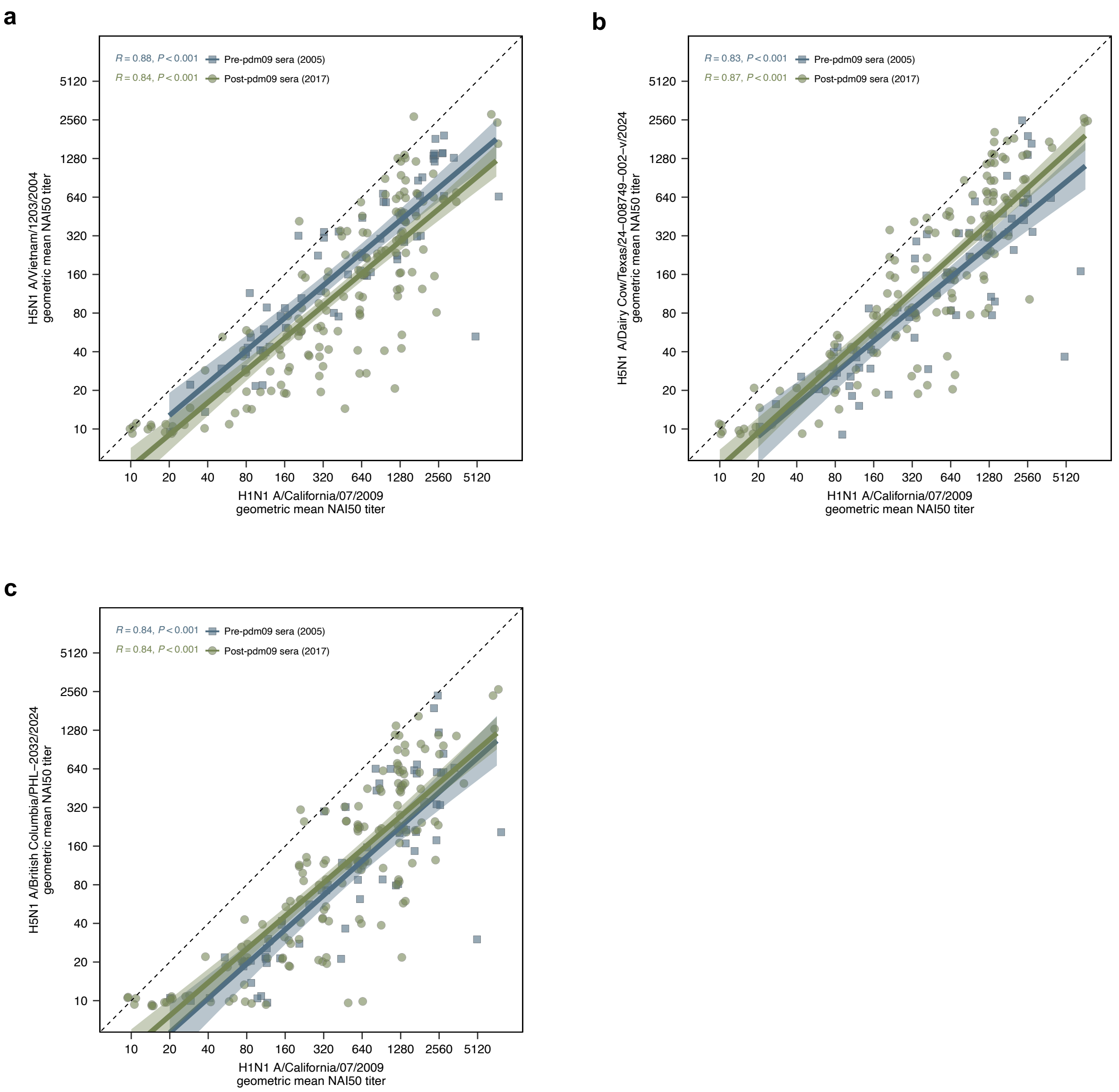


**Extended Data Figure 1. Cross-reactive N1 antibody levels are highly correlated with those against H1N1(pdm09).**

NAI titers against **(a)** H5N1 A/Vietnam/1203/2004, **(b)** A/Dairy Cow/Texas/24-008749-002-v/2024, and **(c)** A/British Columbia/PHL-2032/2024 were plotted versus NAI titers against H1N1(pdm09) A/California/07/2009. Sera samples collected in 2005, pre-H1N1(pdm09) emergence, are shown in blue and those collected in 2017 are shown in green. Correlations between titers for each cohort were determined using Pearson’s correlation coefficient.

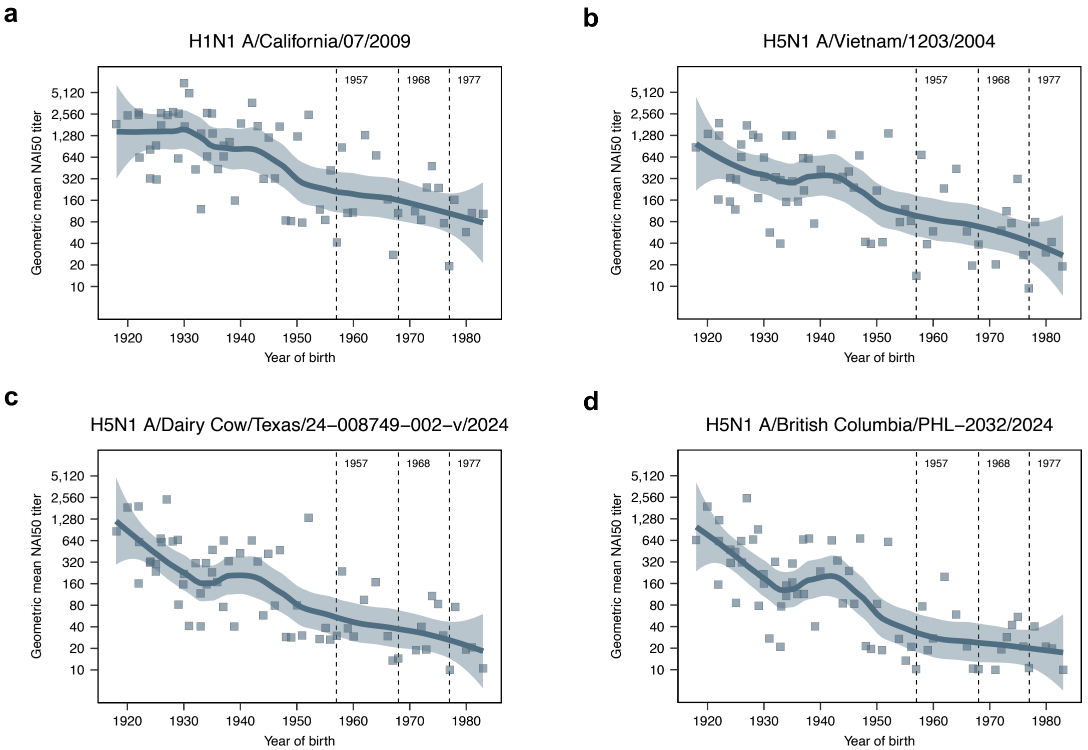


**Extended Data Figure 2. Cross-reactive N1 antibodies were prevalent in the elderly prior to H1N1(pdm09) emergence.**

Banked serum samples from 2005, collected from healthy adults enrolled in vaccine clinical trials prior to vaccination, were assayed for NAI antibodies against H6Nx viruses expressing the NA of **(a)** H1N1 A/California/07/2009, **(b)** H5N1 A/Vietnam/1203/2005, **(c)** H5N1 A/Dairy Cow/Texas/24-008749-002-v/2024, and **(d)** H5N1 A/British Columbia/PHL-2032/2024. Vertical dashed lines represent the emergence of H2N2, H3N2, and H1N1 in 1957, 1968, and 1977, respectively. Each circle represents the geometric mean serum NAI50 titer for a single individual from two to three independent replicates, with locally estimated scatterplot smoothing curves (smoothing parameter = 0.5) and the associated 95% CIs.

### Methods

#### Inclusion and ethics

Experiments using human sera were conducted with the approval of the University of Pennsylvania Institutional Review Board and the Children's Hospital of Philadelphia Institutional Review Board. Serological experiments were completed using de-identified samples. Informed consent was obtained for all individuals.

#### Human serum samples

Sera were collected from adult donors, born from 1927 to 1998, at the Hospital of the University of Pennsylvania during the summer of 2017. Additionally, we obtained de-identified sera from children, born from 2000 to 2016, originally collected at the Children's Hospital of Philadelphia for lead testing in 2017. For pre-H1N1(pdm09) samples, we obtained sera which was collected in 2005 as part of two H5N1 vaccine trials from the National Institutes of Health (ClinicalTrials.gov nos. NCT00115986 and NCT00230750)[1]. For this, we tested a subset of pre-vaccination serum samples from adults, born from 1918 to 1981.

To assess infection-induced antibody responses, we recruited children admitted to the Children's Hospital of Philadelphia during the 2023–24 and 2024–25 influenza seasons who tested positive for influenza A virus infections. Clinical viral isolates were sequenced as described elsewhere[2] to determine the infecting subtype and to confirm that the sequence fell within the circulating viral diversity of that season. Leftover serum samples were collected opportunistically if participants had blood drawn for other reasons. Serum samples collected during acute infection—within seven days of symptom onset—were used to assess pre-existing immunity, while all serum samples collected more than seven days since symptom onset were used to assess infection-induced immunity.

#### Cell lines

293T and 293T–PB1 cells were cultured in Dulbecco's Minimal Essential Medium (DMEM; Corning) supplemented with 10% fetal bovine serum (FBS; Sigma). MDCK–SIAT1 and MDCK–SIAT1–PB1–TMPRSS2 cells were cultured in Minimal Essential Medium (MEM; Corning) with 10% FBS. Co-cultures of these cell lines, used for virus production, were maintained in DMEM with 10% FBS.

**Viruses**

All viruses used in this study were generated by reverse genetics using bi-directional pHW2000 vectors[3]. Reassortant H6Nx viruses were produced with the HA segment of the H6N2 A/Turkey/Massachusetts/3740/1965 strain[4], the NA segment of interest (from H1N1 A/California/07/2009, H5N1 A/Vietnam/1203/2004, H5N1 A/Dairy Cow/Texas/24-008749-002-v/2024, H5N1 A/British Columbia/PHL-2032/2024, or H3N2 A/Thailand/8/2022), and internal genes from A/Puerto Rico/8/1934. For viruses with the A/California/07/2009 NA, reverse genetics plasmids were transfected into a co-culture of 293T and MDCK–SIAT1 cells using Lipofectamine 2000 (Invitrogen). One day post-transfection, cell culture media was replaced with Opti-MEM (Gibco) supplemented with 1 µg/mL tosyl phenylalanyl chloromethyl ketone-treated trypsin (Thermo Scientific), 100 U/mL penicillin (Gibco), and 100 µg/mL streptomycin (Gibco). Transfection supernatants were collected three days post-transfection, then expanded once in embryonated chicken eggs (AVS Bio). As an added biosafety measure, all other H6Nx viruses were generated in a replication-deficient system—in which the PB1 gene is replaced by GFP—which only allows replication in cells expressing PB1[5]. Reverse genetics pHW2000 plasmids (excluding PB1), pHH–PB1flank–eGFP, and pHAGE2–EF1aInt–TMPRSS2–IRES–mCherry were transfected into a co-culture of 293T–CMV–PB1 and MDCK–SIAT1–CMV–PB1–TMPRSS2 cells[5,6], which both constitutively express the PB1 of the A/WSN/1933 strain, using Lipofectamine 2000 (Invitrogen). One day post-transfection, cell culture media was replaced with neutralization assay media (NAM) comprised of Medium-199 (Gibco) with 0.01% FBS

(Sigma), 0.3% bovine serum albumin (BSA; Sigma), 100 U/mL penicillin (Gibco), 100 µg/mL streptomycin (Gibco), 100 µg/mL calcium chloride (Sigma), and 25 mM HEPES (Corning)[7]. Transfection supernatants were collected three days post-transfection, then expanded one to two times on MDCK–SIAT1–CMV–PB1–TMPRSS2 cells in NAM. All viral stocks were clarified by centrifugation and stored at -80°C.

**Enzyme-linked lectin assay (ELLA)**

To titer H6Nx viruses, 96-well Immulon 4HBX extra-high binding flat-bottom plates (Thermo Scientific) were coated with 10 µg/mL fetuin (Sigma) diluted in DPBS (Corning) overnight at 4°C, then washed with PBS (Roche) containing 0.1% Tween-20 (Sigma) (PBS-T) three times using a BioTek 405 LS microplate washer. Two-fold serial dilutions of viruses were prepared in sample buffer comprised of DPBS with calcium and magnesium (Corning) containing 1% BSA (Sigma) and 0.1% Tween-20 (Fisher), then incubated overnight at 37°C on fetuin-coated plates. After 16-18 h, plates were washed three times with PBS-T, then horseradish peroxidase-conjugated peanut agglutinin (PNA-HRP; Sigma), diluted 1:500 in lectin buffer comprised of DPBS with calcium and magnesium (Corning) containing 1% BSA (Sigma), was added and incubated at room temperature for 2 h. Plates were washed three times with PBS-T, then SureBlue TMB substrate (KPL) was added to develop the plates for 5 min, after which 250 mM hydrochloric acid was added to stop the reaction. Plates were read at an optical density (OD) of 450 nm using a SpectraMax 190 plate reader (Molecular Devices). These data were plotted with GraphPad Prism 10, using a nonlinear regression to determine the half maximal effective concentration ($EC_{50}$) for each virus stock.

For NAI assays, two-fold serial dilutions of heat-inactivated serum were prepared in sample buffer and transferred to fetuin-coated plates. An equal volume of H6Nx virus, diluted to 2× its $EC_{50}$ in sample buffer, was added to the sera samples. Additionally, one column of each plate contained sample buffer in place of sera (virus only control) and one column contained neither sera nor virus (background). Plates were incubated for 16-18 h at 37°C, then developed with PNA-HRP and TMB as performed during titering. After background-correcting $OD_{450}$ values, NAI50 titers were determined as the reciprocal of the final serum dilution yielding less than 50% of the mean $OD_{450}$ of virus only control wells. Samples that did not achieve this at the starting dilution of 1:20 were assigned an NAI50 titer of 10.

**Imprinting modeling**

We estimated imprinting probabilities by birth year for samples collected in 2017 from people born in the United States using the *imprinting* R package with default parameters, which implements the approach from Gostic et al.[8,9].

**Statistical analyses**

To test for associations between H1N1(pdm09) NAI titers and H5N1 NAI titers, we calculated Pearson correlation coefficients for each pairing. To test for correlations between NAI titers and different predictors (birth year, HA group 1 imprinting, and N1 imprinting), we calculated Spearman's rank correlation coefficients and computed 95% confidence intervals using Fisher's transformation. We then compared the strengths of these correlations through 1,000 iterations of bootstrapping, calculating the difference in correlation coefficients and computing 95% confidence intervals. We additionally fit linear regressions between NAI titers and each predictor, then compared these models using the Akaike information criterion. To determine if NAI titers were significantly increased following H1N1 or H3N2 infection, we performed paired t-tests between acute and maximum convalescent log2-transformed titers. We corrected *P* values for multiple comparisons within each infection cohort using the Holm–Bonferroni method.

**Data availability**

All data are shown in the main figures and the Extended Data. Source data are provided with this paper.

**Code availability**

Code implementing the imprinting modeling and related statistical analyses is available at https://github.com/HensleyLab-UPENN/N1-imprinting.

**Extended Data References**
